# Clinical outcomes with chronic adaptive versus conventional DBS in Parkinson’s disease: A pilot randomized crossover trial

**DOI:** 10.1101/2025.09.07.25335282

**Authors:** Jun Tanimura, Takehiro Yako, Takao Hashimoto

## Abstract

**Background:** Currently available adaptive deep brain stimulation (aDBS) modulates stimulation guided by real-time beta-band local field potential activity, potentially offering advantages over conventional DBS (cDBS). Clinical evidence under chronic stimulation remains limited.

**Objectives:** To compare aDBS versus cDBS efficacy in patients with Parkinson’s disease (PD) under chronic stimulation.

**Methods:** This was a double-blind, randomized crossover trial in nine PD patients with bilateral subthalamic nucleus DBS. Patients underwent consecutive one-month periods of cDBS and aDBS after a four-month postoperative interval. aDBS used a dual-threshold algorithm adjusting amplitude based on subthalamic beta-band LFP power. Primary outcomes were daily ON/OFF durations and dyskinesia duration under optimized medication. Mixed-effects ANCOVA estimated treatment effects. Bayesian analyses integrated prior evidence to assess probabilities of clinically meaningful differences.

**Results:** No statistically significant differences emerged between treatments. Directional effects varied: ON duration favored cDBS, whereas troublesome dyskinesia and UPDRS scores favored aDBS. Bayesian analyses indicated low probabilities that these differences reached clinical importance (29.4% probability for cDBS advantage on ON time, 7.5% for aDBS advantage on UPDRS Part III). Moderate-to-substantial between-patient heterogeneity was observed in comparative treatment effects across outcomes.

**Conclusions:** The current beta-band LFP-guided aDBS approach with dual-threshold algorithm showed efficacy comparable to cDBS in patients with PD. These findings both call for and provide a basis for appropriately powered follow-up studies to determine whether, for whom, and with which adaptive stimulation strategies aDBS offers greater benefit than cDBS—a key step in the further development of personalized aDBS.

## Introduction

Deep brain stimulation (DBS) of the subthalamic nucleus (STN) is an established treatment for Parkinson’s disease (PD), being particularly beneficial for patients who experience motor fluctuations or medication-induced dyskinesias^1–4^. Conventional DBS (cDBS) provides continuous stimulation at a fixed amplitude, whereas adaptive DBS (aDBS) modulates the stimulation amplitude in real-time, guided by physiological biomarkers such as local field potential (LFP) activity recorded directly from implanted electrodes^5–7^. The aDBS approach, particularly utilizing STN beta-band oscillations peak power amplitude as biomarkers, is based on foundational studies demonstrating their relationship with motor symptoms such as bradykinesia and rigidity^8–12^. The rationale for the use of aDBS is to maintain symptom control while reducing unnecessary stimulation, conserving battery life, and potentially minimizing side effects^13^.

Despite this promising rationale, clinical evidence supporting the use of aDBS over cDBS remains inconclusive^13^. Pioneering studies with well-controlled exploratory designs consistently demonstrated that aDBS resulted in better motor outcomes^14–16^. However, more real-world-oriented follow-up studies, which carefully eliminated perioperative acute-lesion effects or assessed patients under anti-parkinsonian medication, showed no difference in motor outcomes when aDBS was compared to cDBS^17,18^, whereas aDBS was suggested to have advantages in dyskinesia score improvement over cDBS^17^. Recent trials^19–21^ have provided important advances in evaluating chronic aDBS, including extended follow-up periods and diverse trial designs. However, the issue remains unsettled regarding comparative efficacy of aDBS and cDBS due to limited evidence under fully blinded conditions over chronic treatment periods.

In this study, we assessed the clinical efficacy of aDBS versus cDBS in patients with PD using a double-blind, randomized, crossover study design. We used a clinically available dual-threshold aDBS algorithm that continuously adjusted stimulation amplitude according to beta-band LFP activity power in the STN. To ensure assessment under stable chronic conditions, evaluations were performed after a postoperative stabilization period of more than four months and after one month of each stimulation method (in random order) under optimized antiparkinsonian medication.

## Methods

### Study Design and Participants

We conducted a pilot, two-period, double-blind, crossover randomized controlled trial comparing adaptive DBS (aDBS) with conventional DBS (cDBS) in patients with Parkinson’s disease (Figure 1). The trial employed a 1:1 allocation ratio and aimed to assess treatment differences between the two stimulation modes. The analysis was designed to estimate treatment differences between aDBS and cDBS for each outcome.

**Fig. 1.**
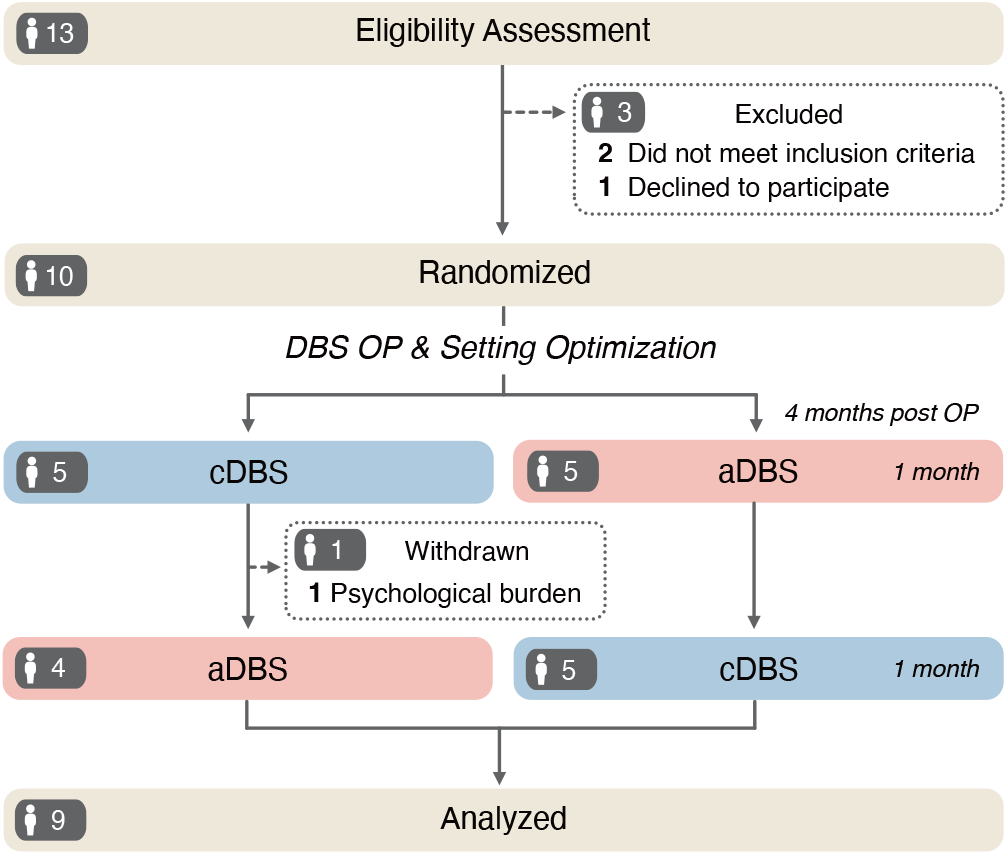
Participant flow: randomized two-period crossover. Flow diagram showing numbers screened, randomized, allocated to each first-period sequence, completed, and analyzed, with exclusions and reasons specified at each stage; recruitment and assessment windows are indicated.

Eligible participants were patients with a clinical diagnosis of Parkinson’s disease, aged under 75 years, with motor complications (motor fluctuations and/or dyskinesia). We excluded patients with cognitive impairment (MMSE < 24), or with significant psychiatric symptoms requiring treatment changes. All participants were recruited from the Movement Disorders Clinic at Aizawa Hospital, Matsumoto, Japan. The study was registered with the UMIN Clinical Trials Registry (UMIN000058937).

### Sample Size

As a pilot study, a formal sample-size calculation for the primary outcomes (ON/OFF time and dyskinesia duration) was not conducted; no previous studies had reported effect-size estimates required for a power calculation at the time we designed this trial^14,15,17,18^. Instead, we referred to secondary motor outcomes that had been evaluated in prior trials. In particular, differences in UPDRS Part III scores between aDBS and cDBS were detected with 8 to 10 study participants with *α* = 0.05^14,15^. Similarly, improvements in dyskinesia scores (Unified Dyskinesia Rating Scale, UDyRS) were detected with approximately 10 participants^18^. Based on these precedents, we aimed to recruit 10–12 participants to evaluate potential treatment differences.

### DBS lead implantation surgery

All patients received standard bilateral STN implantation using the Percept PC system (Medtronic, USA), which enables both cDBS and aDBS. Bilateral lead implantation was performed in a single operative session. We implanted Medtronic 3387 leads under microelectrode guidance with local anesthesia. Prior to surgery, MRI (including heavy T2-weighted, proton, and gradient echo planar T1 sequences with gadolinium enhancement; MAGNETOM Avanto, Siemens Healthineers, Erlangen, Germany) and non-contrast CT scans were obtained with Leksell G Frame to plan target trajectories using Leksell SurgiPlan (Elekta, Stockholm, Sweden). Recording tracks proceeded from anterodorsal to posteroventral at 20–30° from vertical and from lateral to medial at 20° from the sagittal plane. The STN borders were defined physiologically and fitted to atlas maps reconstructed from the Schaltenbrand and Bailey Atlas^22^. Lateral distance to the midline was measured on intraoperative CT scan. We targeted placement of the lowest electrode contact 0–1 mm below the ventral STN border and 10–11 mm lateral to midline, with final electrode locations confirmed by postsurgical MRI. The implantable pulse generator (IPG) was placed subcutaneously in the chest 2–3 days after lead implantation.

### Stimulation setting

In this study, we used a clinically available dual-threshold aDBS algorithm that continuously adjusted stimulation amplitude within a programmed range according to beta-band LFP activity power in the STN, following the Clinician Programmer Guide^23,24^. Immediately after surgery, we observed LFP with stimulation OFF to define (1) a contact pair to sense the large beta-band peak, which stands out sharply from the background activity, (2) a stimulating contact for monopolar stimulation, and (3) the range of LFP (Hz) to track (fixed to 5 Hz range). We targeted a peak within 8–30 Hz with a peak amplitude of *>* 1.2 *µ*V^23,24^. We searched for suitable sensing contacts bilaterally and finally chose one ipsilateral contact pair for the bilateral control. Using the stimulating contact, which is defined as the flanked contact with the pair of sensing contacts, cDBS stimulation settings were then configured with the default pulse duration of 60 *µ*s and frequency of 130 Hz; the stimulation amplitude was initially determined by gradually increasing the amplitude day-by-day and was set just below that producing persistent adverse effects. Patients received cDBS therapy for the first three months after surgery.

During the fourth month after surgery, which was one month prior to randomization, we configured the remaining parameters for aDBS and tested tolerance for aDBS stimulation over that month. The previously defined sensing contacts were reconfirmed in all patients, with no changes required. Next, LFP threshold values were configured for the Dual Threshold Mode. During the process, the stimulation current was used as an actuator to capture two LFP-power thresholds^23,24^: first, patients received the highest amplitude just below that producing persistent adverse effects (upper therapeutic limit current), and the observed beta power during this stimulation was defined as the lower LFP threshold; subsequently, patients received a lower amplitude (fixed at 20% below the upper therapeutic limit current), and the observed beta power during this stimulation was defined as the upper LFP threshold. (Note that this latter approach for defining the upper LFP threshold deviates from the guidance^23,24^; this fixed 20% reduction—rather than using a level guided by the minimal current needed for symptom improvement—was adopted because, in our experience, it is often difficult to determine the threshold for the initial appearance of parkinsonian symptom improvement reliably.)

Finally, we defined the stimulation setting for aDBS: the higher stimulation current level was set as the upper therapeutic limit used during the LFP-threshold determination process, and the lower stimulation current level was fixed at 20% below the higher level. The ramp time, which is the time of changing between the two stimulation amplitudes, was set to 2.5 min for the upward direction and 5 min for the downward direction. After setting the aDBS program, the setting for cDBS current amplitude was also updated to use the same upper therapeutic limit. The patients entered the randomized crossover periods after confirmation of stable condition for a one-month aDBS period. During the one-month pre-randomization aDBS period, patients underwent 1–2 additional adjustment sessions to optimize the parameters. Throughout the titration periods, no changes to sensing or stimulation contacts were required. Beta power threshold crossings, confirming appropriate closed-loop operation, were observed in all patients during the initial configuration and subsequent titration sessions.

### Randomization and Integrity of the Blinding

An independent statistician prepared 12 opaque, sealed envelopes containing treatment sequence assignments. Four months after DBS implantation, personnel not involved in patient assessment or data analysis randomly selected and opened one envelope to determine the treatment sequence for each participant. Participants were assigned to receive either cDBS followed by aDBS (Sequence 1) or aDBS followed by cDBS (Sequence 2), with each treatment period lasting one month (Figure 1). Both participants and outcome assessors remained blinded throughout the study. Device programming was performed by independent, unblinded personnel who were not involved in outcome assessments. All analyses were conducted only after completion of all blinded assessments. Participants were not systematically asked which stimulation mode they believed they received.

### Outcomes

Clinical outcomes were assessed at the end of each one-month treatment period. Baseline (“preoperative”) values for all outcomes were obtained during an inpatient admission within 7 days prior to DBS implantation. Primary outcomes were daily ON time, OFF time, and dyskinesia duration (troublesome and non-troublesome), based on diary records averaged over the 7-day diary period. Secondary outcomes included total UPDRS score, individual UPDRS subscales (Parts I, II, III, IV), Mini-Mental State Examination (MMSE), Parkinson’s Disease Questionnaire-39 (PDQ-39), and Schwab and England Activities of Daily Living (ADL) Scale. All UPDRS assessments during crossover phases were performed in the medication-ON state with optimized medication.

### Statistical Analysis

Treatment effects were estimated using mixed-effects ANCOVA models including treatment, period, sequence, and the standardized preoperative value of the corresponding outcome as fixed effects, with a participant-specific random intercept. Results are reported as adjusted mean differences (aDBS minus cDBS), 95% confidence intervals, and standardized effect sizes (Cohen’s d). Two-sided p-values were adjusted using the Benjamini–Hochberg method. Differential carryover was assessed using treatment-by-period interactions; missing data were analyzed using available observations without imputation. Between-patient heterogeneity was summarized using intraclass correlation coefficient (ICC). Robustness was assessed using reduced ANCOVA and unadjusted paired-comparison models. For ON time and UPDRS Part III, complementary Bayesian analyses used literature-informed and weakly informative priors to estimate the probabilities of clinically meaningful differences and comparable effectiveness based on established minimal clinically important differences (MCID). Analyses were performed using R version 4.3.1. Full model specifications, effect-size calculations, sensitivity analyses, prior distributions, MCIDs, and MCMC procedures are provided in Supplementary Methods S1. Methods and results for the post hoc exploratory treatment-by-baseline interaction analyses are provided in Supplementary Methods S2.

### Study Registration and Ethics

The study was approved by the Ethics Committee of Aizawa Hospital (Approval No. 2020-064: R2-37) and conducted in accordance with the Declaration of Helsinki. All participants provided written informed consent.

## Results

### Participants and Baseline Characteristics

Of 10 patients initially enrolled between January 2022 and January 2025, one withdrew due to psychological burden related to participation, leaving 9 patients who completed both randomized periods and were included in the final analysis (Figure 1). Participants were randomized to two sequences:

Group 1 (n=5) received cDBS in Phase 1 followed by aDBS in Phase 2, while Group 2 (n=4) received aDBS in Phase 1 followed by cDBS in Phase 2. All 9 participants adhered to their assigned stimulation protocols throughout both one-month periods without protocol deviation. For each outcome, we analyzed all available observations (18 per measure, except MMSE with 15 observations).

Baseline characteristics are summarized in Table 1. The median age was 62 years (IQR 54–65) with a median disease duration of 10 years (IQR 8–11). Four participants (44.4%) were female. The median LEDD was 932 mg/day (IQR 881–1182), and the Hoehn & Yahr stage was 3 in the ON state (IQR 1–3) and 4 in the OFF state (IQR 4–4). Patients showed moderate to severe motor impairments, with median UPDRS Part III scores of 37 in the OFF state and 15 in the ON state. Motor complications were present in all participants (UPDRS Part IV median 8). Baseline time outcomes showed median daily ON time of 9.7 hours (IQR 8.0–10.1), OFF time of 8.3 hours (IQR 7.9–10.0), troublesome dyskinesia time of 0.3 hours (IQR 0.1–1.7), and non-troublesome dyskinesia time of 1.8 hours (IQR 1.0–2.4).

**Table 1.** Baseline characteristics of participants. Baseline demographic and clinical characteristics of the study participants at the preoperative assessment (n=9). UPDRS: Unified Parkinson’s Disease Rating Scale; H&Y: Hoehn & Yahr stage; IQR: interquartile range; MMSE: Mini-Mental State Examination; PDQ-39: Parkinson’s Disease Questionnaire; LEDD: levodopa-equivalent daily dose.

| Variable | Median [IQR] or N (%) |
| --- | --- |
| Female, n (%) | 4 (44.4%) |
| Age, y | 62 [54–65] |
| Disease duration, y | 10 [8–11] |
| LEDD, mg/day | 932 [881–1182] |
| ADL (Schwab & England) | 90 [90–100] |
| Hoehn & Yahr, ON/OFF | 3 [1–3] / 4 [4–4] |
| ON time, h/day | 9.7 [8.0–10.1] |
| OFF time, h/day | 8.3 [7.9–10.0] |
| Dyskinesia time, h/day |  |
| Troublesome | 0.3 [0.1–1.7] |
| Non-troublesome | 1.8 [1.0–2.4] |
| UPDRS |  |
| Part I | 1 [0–2] |
| Part II, ON/OFF | 3 [1–7] / 18 [17–27] |
| Part III, ON/OFF | 15 [3–18] / 37 [30–47] |
| Part IV | 8 [7–10] |
| Total, ON/OFF | 26 [13–34] / 69 [56–86] |
| MMSE | 29 [28–30] |
| PDQ-39 | 45 [30–75] |

### Comparison of aDBS with cDBS

Figure 2 shows the distribution of clinical outcomes across preoperative, cDBS, and aDBS conditions. No differential carryover was detected for any outcome using likelihood-ratio tests (all p *≥* 0.105, Supplementary Table S1).

**Fig. 2.**
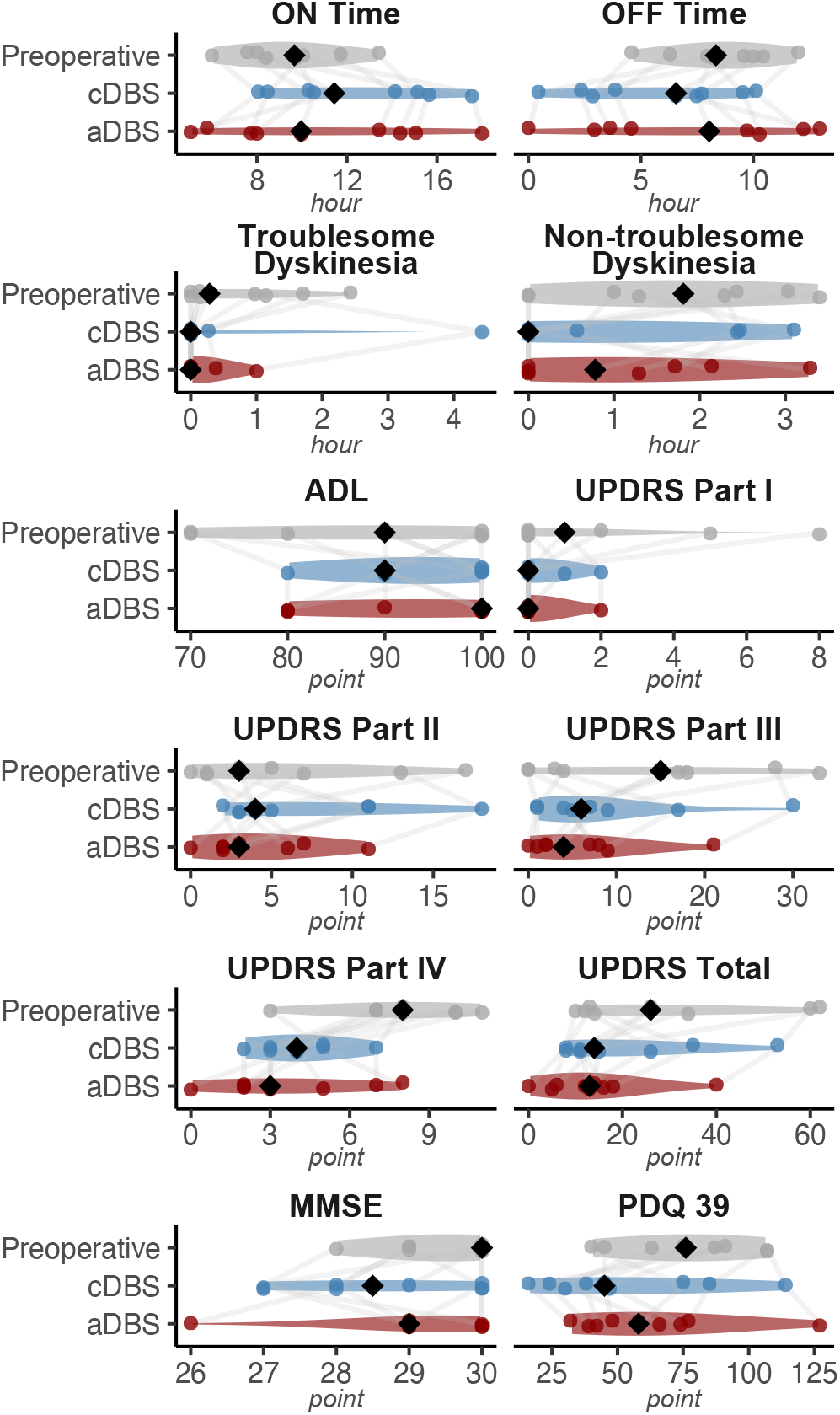
Distribution of clinical outcomes across study conditions. For each outcome, distributions at Preoperative, cDBS, and aDBS are shown (violin plots). Data from both randomization groups are pooled. Individual points are overlaid, and black diamonds indicate the median.

The comparison of aDBS versus cDBS revealed heterogeneous treatment effect directions across clinical domains, with no statistically significant differences (Figure 3). The ON time differed between treatments by 1.91 hours/day favoring cDBS (d = −0.94), which did not reach the minimal clinically important difference (MCID) of *±*2 hours/day. Likewise, the OFF-time differed between treatments by 0.73 hours/day favoring cDBS (d = −0.74). The duration of troublesome dyskinesias showed a small change favoring aDBS (−0.10 hours per day, d = 0.35). Both cDBS and aDBS reduced the total UPDRS score, with the reduction by aDBS exceeding that of cDBS (−5.93 points, d = 1.20). However, this difference did not reach the 8-point minimal clinically important difference^25^. The reduction in UPDRS Part III scores also showed a larger reduction in the aDBS stimulation periods compared to the cDBS stimulation periods (−2.68 points, d = 0.87), which did not reach the MCID of −5 points^25,26^. In summary, no statistically significant differences emerged across all outcomes, and treatment differences remained below minimal clinically important difference thresholds for outcomes with established MCIDs (ON time, UPDRS scores), suggesting similarity in population-level clinical effectiveness between aDBS and cDBS.

**Fig. 3.**
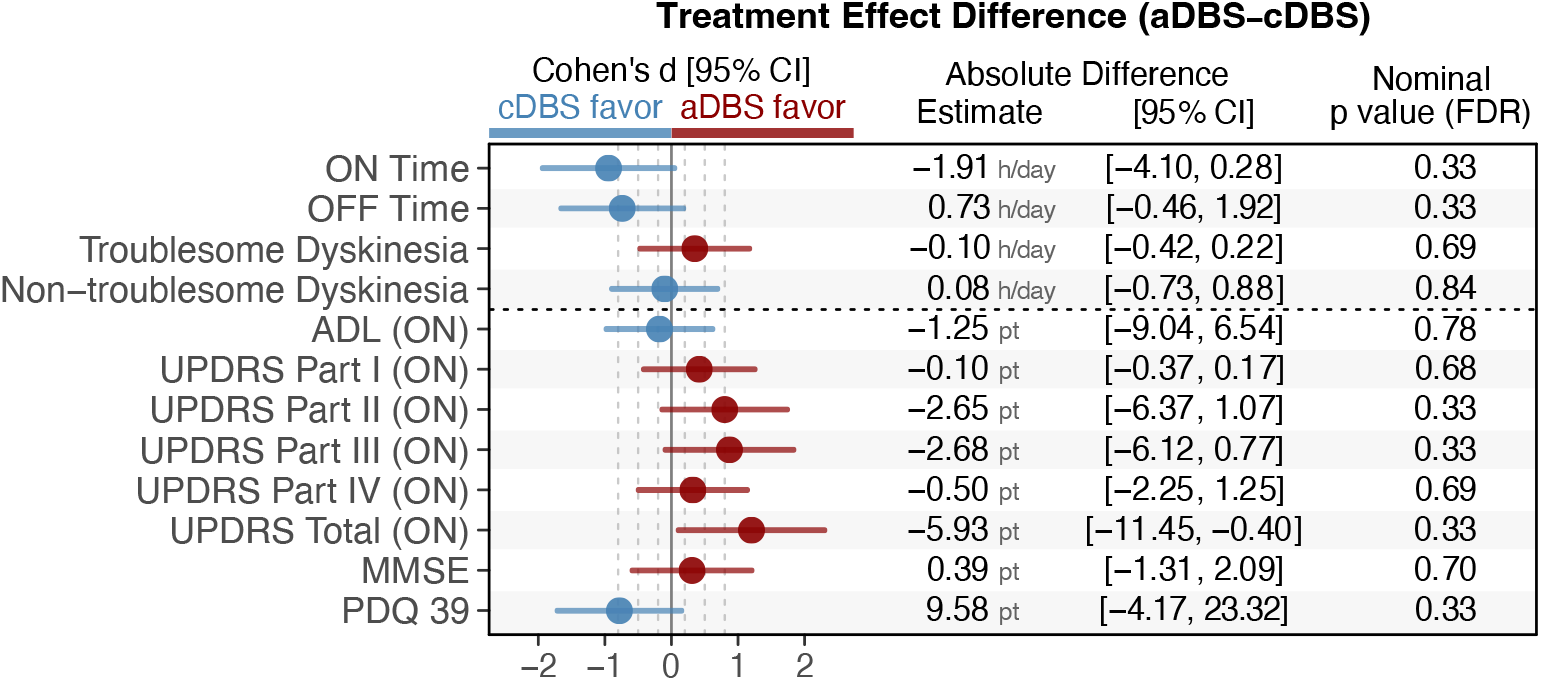
Treatment effects comparing aDBS and cDBS. Forest plot of treatment effect sizes (Cohen’s d) with 95% confidence intervals. Blue indicates cDBS advantage; red indicates aDBS advantage. Gray dashed lines show conventional thresholds (0.2, 0.5, 0.8). Table displays absolute differences.

Sensitivity analyses demonstrated high consistency with the primary findings in terms of the direction and magnitude of treatment effects across all outcomes (Supplementary Table S2 and Supplementary Figure S1).

### Bayesian Complementary Findings

For ON time and UPDRS Part III, we conducted Bayesian analyses integrating prior evidence from recent trials with current data, providing intuitive clinical interpretations (Figure 4, Supplementary Table S3).

**Fig. 4.**
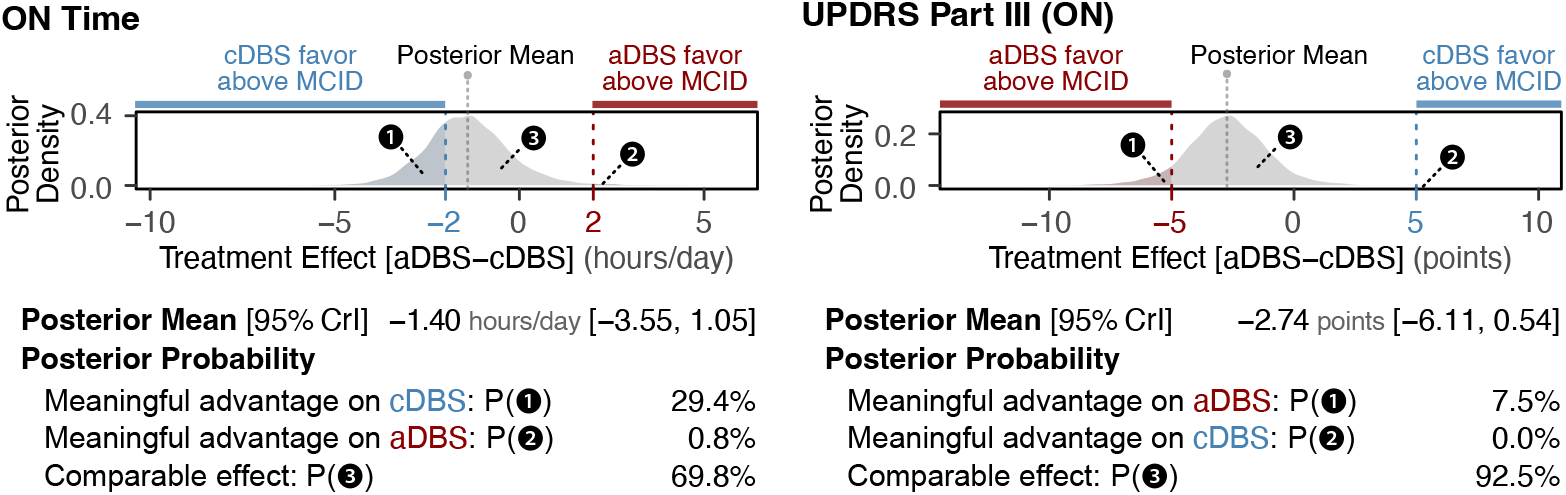
Treatment effect probabilities with clinical thresholds. Posterior probability distributions for treatment effects (aDBS–cDBS) from Bayesian mixed-effects crossover models. Dashed lines indicate minimal clinically important difference (MCID) thresholds (*±*2 hours/day for ON time, *±*5 points for UPDRS Part III). CrI: credible interval.

The Bayesian estimate of the treatment effect (posterior mean of aDBS minus cDBS) was −1.40 hours/day (95% CrI: −3.55 to 1.05) for ON time and −2.74 points (95% CrI: −6.11 to 0.54) for UPDRS Part III, maintaining the directional patterns observed in the frequentist estimates. Probabilities of clinically meaningful differences in these directions were low for both ON time (29.4% for cDBS advantage exceeding −2 hours/day MCID) and UPDRS Part III (7.5% for aDBS advantage exceeding −5 points MCID). Probabilities of clinically comparable treatment effectiveness (difference within *±* MCID) were high for both ON time (69.8%) and UPDRS-III (92.5%), together suggesting similarity in population-level clinical effectiveness. Sensitivity analyses demonstrated that posterior estimates appropriately responded to prior specifications while maintaining consistent directional conclusions (Supplementary Table S3).

### Between-Patient Heterogeneity

We summarized patient-level heterogeneity in the treatment differences (aDBS minus cDBS) using the intraclass correlation coefficient (ICC) from the primary mixed-effects models. The median ICC across outcomes was 0.67 (IQR, 0.50– 0.86), indicating moderate-to-substantial between-patient heterogeneity; approximately 67% of the remaining variance in treatment differences was attributable to differences between patients rather than within-patient variation (Supplementary Table S4).

Exploratory, hypothesis-generating analyses examining whether baseline characteristics modified treatment differences are presented in the Supplementary Material (Supplementary Methods S2, Supplementary Figure S3, and Supplementary Table S5), with the complete interaction estimates and leave-one-out results available in the data repository.

### Adverse Events

During the assessment after each one-month treatment period, none of the patients reported adverse events (such as paresthesia, dysarthria, dysphagia etc.). Furthermore, no signs of device-related infections, cerebral hemorrhage, or seizures were observed or reported.

## Discussion

### Key findings

In this pilot, double-blind crossover trial, we evaluated the relative effectiveness of a current beta-band aDBS approach using subthalamic local field potential activity for adaptive stimulation. We observed no statistically significant differences between aDBS and cDBS. Directional patterns emerged: ON times tended to be longer and OFF times shorter with cDBS, whereas aDBS showed trends toward benefits for troublesome dyskinesias and UPDRS scores. Bayesian analyses integrating prior evidence indicated low probabilities of clinically meaningful differences for these directions (29.4% for cDBS advantage on ON time, 7.5% for aDBS advantage on UPDRS Part III), suggesting similarity in population-level effectiveness between the two DBS modes. Moderate-to-substantial between-patient heterogeneity in treatment differences was observed across outcomes.

### Comparison with Recent Literature

ADAPT-PD^19^ is the largest trial to date using the same Percept PC adaptive DBS system, and its prespecified primary endpoint has also established the comparable efficacy of dual-threshold aDBS to cDBS. In its exploratory comparison, aDBS was associated with a longer ON time (13.62 vs 12.30 h/day; difference, +1.3 h/day) and a lower MDS-UPDRS Part III score (24.0 vs 26.5; difference, −2.6 points) than cDBS; the ON-time difference reached the defined 1hour minimal clinically important change (MCIC), whereas the UPDRS Part III difference remained below the corresponding MCID (3.25 points), and the authors emphasize that confirmatory inference from these comparisons is precluded. Our study reached the same conclusion: in a randomized, double-blind framework, with more conservative MCID thresholds (*±* 2 h/day for ON time; *±* 5 points for UPDRS Part III), neither outcome reached a clinically meaningful difference. The adjusted ON-time difference (−1.91 h/day; raw means, 10.82 vs 12.36 h/day) and the UPDRS Part III difference (−2.68 points favoring aDBS) corresponded to low Bayesian-estimated probabilities of a clinically meaningful advantage for either mode, although the ON-time estimate was directionally opposite to ADAPT-PD.

The two cohorts were similar in age and both had longstanding PD, and absolute ON time during cDBS was nearly identical (12.30 h/day in ADAPT-PD vs 12.36 in our study; Supplementary Figure S2), indicating that both cohorts were optimized to a comparable cDBS standard^4,34^. Against this background, the opposite ON-time direction is best read as a design- and population-sensitive signal of modest clinical weight. The recruitment designs differed: our participants were evaluated soon after STN-DBS implantation, whereas ADAPT-PD enrolled patients already on stable chronic cDBS for a mean of 3.4 years; whether longer DBS experience relates to greater ON-time gains at aDBS initiation, a question explicitly raised in ADAPT-PD, remains unresolved and will require replication. The open-label design of ADAPT-PD, as the authors note, may inflate the apparent aDBS benefit through placebo effects, and its flexible optimization, with adjustments permitted over a period of up to 60 days, also differs from our prespecified, double-blind protocol. Substantial between-patient heterogeneity could also move group-level estimates. We therefore regard the two trials, with their differing designs, as complementary: ADAPT-PD illustrates the clinical potential of aDBS under a flexible, better-powered design, whereas our pilot study supports the robustness of comparable effectiveness between the two stimulation modes under controlled, unbiased conditions.

In another chronic aDBS study, Busch et al.^21^ reported real-world outcomes in 8 patients using the same Percept PC device in an open-label design over four months. Compared to cDBS, aDBS was associated with improved overall well-being and a non-significant trend toward enhanced general movement; 6 of 8 patients elected to continue aDBS. Taken together with ADAPT-PD and our trial, objective motor outcomes appear comparable between modes across the chronic aDBS literature, whereas patient-reported measures such as well-being and long-term preference tend to favor aDBS.

Finally, the ADAPT-PD also compared dual-threshold (DT) and single-threshold (ST) aDBS algorithms in a randomized design and found no difference in the primary outcome, with exploratory analyses suggested somewhat greater improvements with DT than with ST relative to cDBS. These findings support our decision to focus on the DT algorithm in the present study.

We observed moderate-to-substantial between-patient heterogeneity in aDBS minus cDBS treatment differences. Emura et al.^27^ found that baseline clinical characteristics were associated with differential treatment responses to aDBS versus cDBS. Whether such heterogeneity is patient-intrinsic, or can instead be modified and tailored through personalized stimulation, remains an open question.

### Limitations and Generalizability

As a pilot study, the lack of formal sample size estimation for primary outcomes potentially limits statistical power. We did not ask participants to guess their stimulation mode during the blinded periods, which would have further supported effective blinding. Each stimulation mode was blindly evaluated over a one-month period; although this duration exceeds that of other studies, it still limits inferences regarding further long-term efficacy or durability of treatment effects over months to years.

Contact selection prioritized biomarker signal quality on sensing contacts, which may have constrained stimulation contact optimization equally across both aDBS and cDBS modes. Unilateral sensing was used for bilateral stimulation control: although bilateral functional connectivity between the STNs is statistically detectable, its magnitude is relatively small^33^, and whether unilateral sensing yields adequate clinical outcomes remains an open question. Future studies should systematically compare unilateral and bilateral sensing strategies. The beta-band peak and its correspondence with clinical symptoms were not re-confirmed chronically after randomization, which may limit our interpretation in terms of biomarker stability. Systematic stimulation pattern data such as time spent at upper and lower amplitude limits and stimulation adaptation frequency were not prospectively collected during the crossover periods, limiting our ability to quantify the degree of aDBS adaptation. Future trials should include continuous recording of these metrics.

Our study used specific hardware (Percept PC) and LFP-based sensing with beta-band amplitude thresholding; thus, findings should be extrapolated cautiously to other biomarkers, algorithms, or devices. Device-level metrics (total energy delivered, battery consumption) and some patient-centered outcomes such as sleep quality, non-motor symptoms, and patient preference for the two DBS modes were not systematically assessed during each stimulation period.

The present findings should be interpreted as applying to the current form of beta-band LFP-guided aDBS with the specific programming strategy used in this study, rather than to aDBS as a whole. Alternative adaptive strategies using other neural features, such as beta bursts in the dorsolateral STN or gamma oscillations in motor cortex^28–32^, may yield different clinical profiles and should be evaluated in future comparative studies. Being a single-center study in Japan, external validity may be limited for settings with different healthcare systems, patient populations, or clinical practices in DBS management; thus, results should be interpreted with reference to our baseline patient characteristics and DBS programming methodology.

In conclusion, this pilot, randomized crossover trial demonstrated that the current beta-band LFP-guided aDBS approach with a dual-threshold algorithm showed comparable population-level efficacy to cDBS in real-world clinical settings for patients with Parkinson’s disease. These findings both call for and provide a basis for appropriately powered follow-up studies to determine whether, for whom, and with which adaptive stimulation strategies aDBS offers greater benefit than cDBS—a key step in the further development of personalized aDBS.

## Data Availability

All data produced in the present study are available upon reasonable request to the authors.

https://doi.org/10.7910/DVN/U1SNWI

## ACKNOWLEDGEMENTS

The authors thank Dr. Shoji Yomo for his contributions to surgical planning using the SurgiPlan system, and Dr. Thomas Wichmann for his critical and constructive comments on the manuscript.

## DATA AND CODE AVAILABILITY

All data and code are available through Harvard Dataverse (DOI:10.7910/DVN/U1SNWI).

## GENERATIVE AI USAGE

The authors used ChatGPT (OpenAI), Claude (Anthropic), and Gemini (Google) to improve readability, grammar, word choice, and flow. After using these tools, the authors carefully reviewed and edited the content as needed and take full responsibility for the content of the published article.

## AUTHOR CONTRIBUTIONS

**Research project:** A. Conception; B. Organization; C. Execution.

**Statistical Analysis:** A. Design; B. Execution; C. Review and Critique.

**Manuscript Preparation:** A. Writing of the first draft; B. Review and Critique.

**JT:** 1A, 1B, 1C, 2A, 2B, 3A, 3B.

**TY:** 1C, 3B.

**TH:** 1A, 1B, 2A, 2B, 2C, 3B.

## Ethical Compliance Statement

### Institutional review board approval

The study was approved by the Ethics Committee of Aizawa Hospital (Approval No. 2020-064: R2-37) and conducted in accordance with the Declaration of Helsinki.

### Patient consent

Written informed consent was obtained from all participants prior to any study procedures and documented in the study records.

### Ethical publication

We confirm that we have read the Journal’s position on issues involved in ethical publication and affirm that this work is consistent with those guidelines.

