## Supplemental Material for "Clinical outcomes with chronic adaptive versus conventional DBS in Parkinson’s disease: A pilot randomized crossover trial"

### Contents

- **Supplementary Methods S1**. Primary, sensitivity, and Bayesian statistical analyses
- **Supplementary Methods S2**. Post hoc exploratory treatment-by-baseline interaction analyses
- **Supplementary Results A**. Exploratory treatment-by-baseline interaction analyses
    - **Supplementary Figure S3**. Post hoc exploratory treatment-by-baseline interaction analysis.
    - **Supplementary Table S5**. Direction of leave-one-out sign-stable treatment-by-baseline interaction estimates (|r| ≥ 0.5).
- **Supplementary Results B**. Additional sensitivity and cross-study analyses
    - **Supplementary Figure S1**. Comparison of effect sizes across model specifications
    - **Supplementary Figure S2**. Cross-study comparison of treatment-phase ON time

### Supplementary Methods S1. Primary, sensitivity, and Bayesian statistical analyses

*Mixed-effects Analysis*

We compared the efficacy of aDBS with that of cDBS. We used a mixed-effects ANCOVA model with fixed effects for treatment, period, sequence, and standardized baseline values, and subject-specific random intercepts, thereby explicitly modeling and quantifying between-patient variability while adjusting for design-related period and sequence effects, as well as baseline differences in the outcome measure assessed prior to surgery[S1-S3].

Treatment differences (aDBS minus cDBS) were estimated in original units as adjusted mean differences with 95% confidence intervals (CIs), accompanied by standardized effect sizes (Cohen's d). Cohen's d was calculated as the treatment difference estimate divided by the model residual standard deviation, which represents within-patient variability estimated from the crossover design, enabling interpretation of effect size as the magnitude relative to within-patient variation. Confidence intervals for treatment estimates and Cohen's d were constructed using t-distribution based on model degrees of freedom. All tests were two-sided with p-values adjusted for multiple comparisons using the Benjamini-Hochberg method. Time-based outcomes were analyzed on the log(1+x) scale to handle zero values, with back-transformation of marginal estimates for presentation on the original time scale. We tested for differential carryover effects by comparing mixed models with and without a treatment-by-period interaction using likelihood-ratio tests (LRTs)[S1, S4, S5]. Missing data were handled through available case analysis without imputation.

For clinical interpretation, we defined minimal clinically important differences (MCIDs) based on established thresholds: ±2 hours/day for ON time[19, 20]; ±5 points for UPDRS Part III[25, 26]; ±8 points for UPDRS Total[25].

We quantified between-patient heterogeneity using the intraclass correlation coefficient (ICC)[S1, S6], derived from the random intercept variance component of the mixed-effects ANCOVA model in the primary analysis. The ICC was calculated as $ICC = \sigma_b^2/(\sigma_b^2 + \sigma^2)$, where $\sigma_b^2$ represents the between-patient variance and $\sigma^2$ the residual variance.

*Sensitivity Analyses*

To assess the robustness of our primary analysis, we conducted sensitivity analyses with two additional models: 1) a reduced ANCOVA model that excluded crossover-related terms (Period and Sequence); 2) a crude paired comparison that computed treatment differences (aDBS minus cDBS) without adjustment. For both sensitivity models, we calculated treatment effect estimates and Cohen's d effect sizes with 95% confidence intervals using identical methods as the full primary analysis.

*Bayesian Complementary Analysis*

For ON time and UPDRS Part III, we conducted Bayesian analyses integrating prior evidence from recent chronic aDBS trials with current data, enabling intuitive clinical interpretation through probabilistic assessment of treatment effects. These outcomes were selected for Bayesian analysis because (1) prior information on treatment difference (aDBS minus cDBS) was available from recent chronic aDBS trials[19, 20], enabling construction of literature-informed prior distributions, and (2) established MCID thresholds exist for both outcomes[19, 20, 25, 26]. We used the same model structure as the primary mixed-effects ANCOVA, estimating posterior distributions of treatment effects using Markov Chain Monte Carlo (MCMC) sampling implemented in the brms package (version 2.21.0) in R.

We specified two prior distributions for the treatment effect coefficient (aDBS minus cDBS) as a sensitivity analysis. (1) Literature-informed priors were constructed as normal distributions based on recent chronic aDBS trials[19, 20]: for ON time, prior = Normal(1.5, 3.0); for UPDRS Part III, prior = Normal(-3, 4). These priors reflect expectations of advantages of aDBS over cDBS on these outcomes based on prior evidence. Moderately wide standard deviations were used to appropriately incorporate this external evidence given methodological heterogeneity across trials (different device[20] or open-label design[19]). (2) Weakly informative priors used normal distributions centered at zero with moderate standard deviations (Normal(0, 4) for ON time; Normal(0, 6) for UPDRS Part III), representing minimal prior knowledge while reflecting realistic effect ranges. All other model parameters used default brms priors. For each outcome and prior specification, we ran 4 independent MCMC chains with 4000 iterations each (2000 warmup iterations discarded). Convergence was assessed using R-hat statistics (<1.01 indicating convergence) and effective sample sizes (all >3400, indicating adequate precision for posterior inference).

We calculated: (1) posterior means and 95% credible intervals (CrIs), representing the range containing 95% of the posterior probability mass; (2) probabilities of clinically meaningful effects, defined as the probability that treatment differences exceed established MCID thresholds in each direction (favoring aDBS or cDBS); and (3) probabilities of comparable treatment effectiveness, defined as the probability that treatment differences fall within ±MCID.

Analyses were performed using R version 4.3.1 with relevant packages including lme4, lmerTest, emmeans, and brms.

### Supplementary Methods S2. Post hoc exploratory treatment-by-baseline interaction analyses

As a post hoc, hypothesis-generating analysis, we examined whether baseline characteristics modified the relative effect of aDBS versus cDBS. For each outcome–modifier combination, we fitted the mixed-effects ANCOVA model that included treatment, period, sequence, the standardized baseline value of the corresponding outcome, the baseline modifier, the treatment-by-modifier interaction, and a participant-specific random intercept.

Given the sample size (n=9) and the 192 outcome–modifier combinations, all analyses are exploratory and are not intended to support treatment-selection recommendations.

Time-based outcomes were analyzed after log(1+x) transformation, consistent with the primary analysis. Continuous baseline modifiers were standardized and sign-aligned so that higher values represented greater baseline burden. To provide a scale-free descriptive measure, model-based correlation coefficients were calculated from the interaction t statistic as $r = t/\sqrt{t^2+df}$. The outcome direction was also sign-aligned so that positive r values indicated a relative aDBS direction and negative values a relative cDBS direction with higher baseline burden. No multiplicity correction or confirmatory hypothesis testing was applied.

To assess sensitivity, we repeated each interaction analysis after sequentially excluding each participant. An estimate was classified as leave-one-out sign-stable when its direction did not reverse in any iteration. This classification was used only as a descriptive sensitivity assessment and was not interpreted as evidence of statistical robustness.

### Supplementary Results A

#### Exploratory treatment-by-baseline interaction analyses

The exploratory analysis evaluated 192 outcome–modifier combinations. Sign-stable estimates are displayed in Supplementary Figure S3, and a descriptive summary in Supplementary Table S5. The complete interaction estimates and leave-one-out results are available in the Harvard Dataverse repository (DOI: 10.7910/DVN/U1SNWI). These findings should be interpreted solely as hypotheses for future, adequately powered studies.

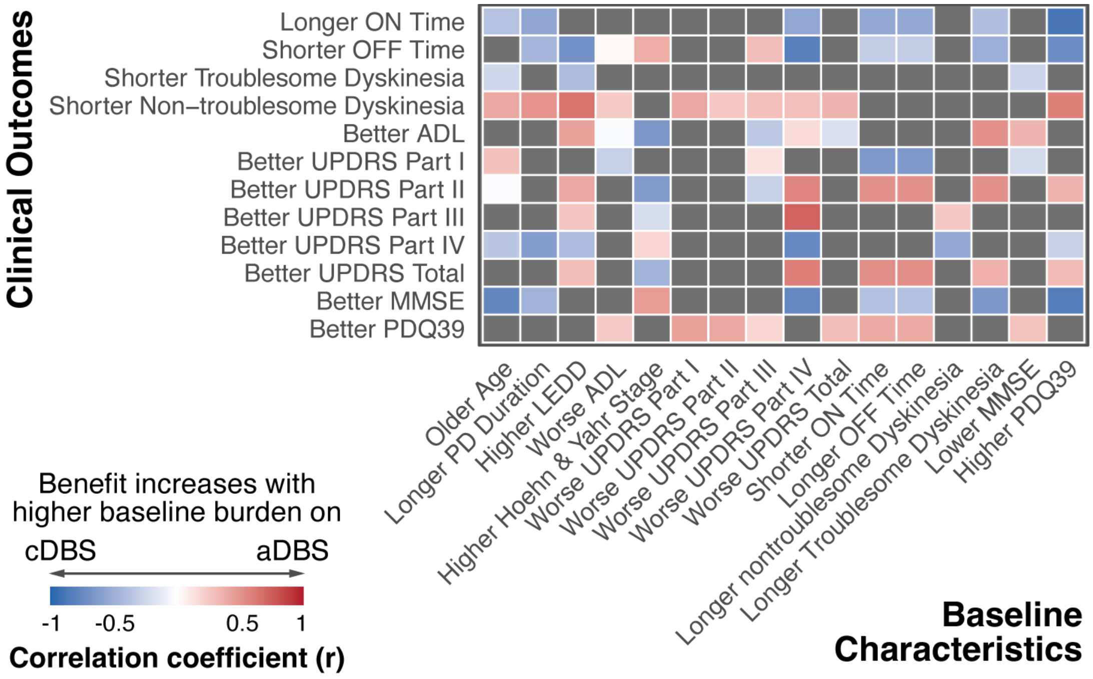


**Supplementary Figure S3. Post hoc exploratory treatment-by-baseline interaction analysis.** Cells display model-based correlation coefficients (r; sign convention as in Supplementary Methods). Gray cells indicate estimates that reversed direction in at least one leave-one-out iteration. The analysis is exploratory and hypothesis-generating.

**Supplementary Table S5. Direction of leave-one-out sign-stable treatment-by-baseline interaction estimates (|r| ≥ 0.5).** r < 0 and r > 0 denote the cDBS and aDBS directions, respectively (Supplementary Methods). Directions describe the sign of exploratory estimates and are not treatment recommendations. UPDRS: Unified Parkinson's Disease Rating Scale; PDQ-39: Parkinson's Disease Questionnaire; LEDD: Levodopa-equivalent daily dose.

| Outcome domain | Interaction direction (higher baseline burden) | Baseline modifiers |
|---|---|---|
| Motor fluctuations (ON/OFF time) | toward cDBS (r < 0) | PD duration, LEDD, UPDRS-IV, PDQ-39 |
| Motor severity (UPDRS-III) | toward aDBS (r > 0) | UPDRS-IV |

### Supplementary Results B. Additional sensitivity and cross-study analyses

Supplementary Figures S1 and S2 present additional sensitivity and cross-study analyses referenced in the main manuscript.

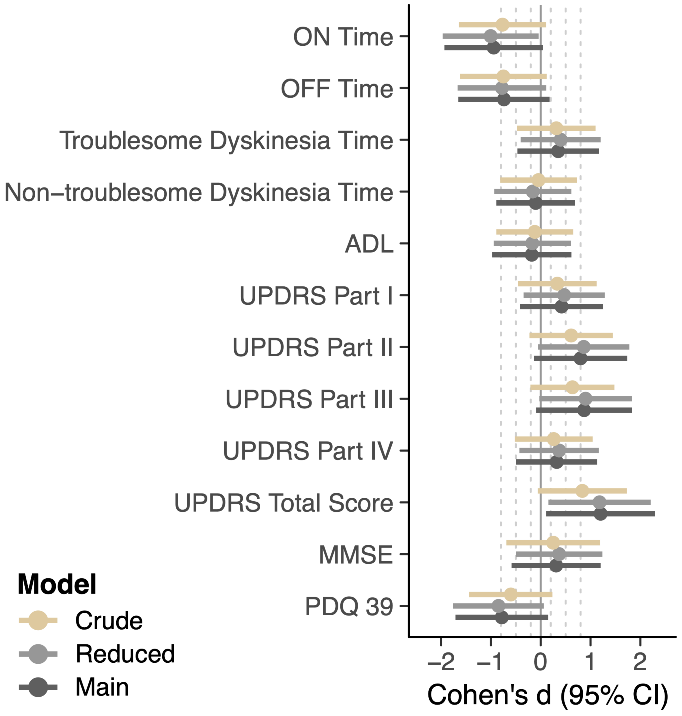


**Supplementary Figure S1. Comparison of effect sizes across model specifications**

Forest plot comparing Cohen's d effect sizes (with 95% confidence intervals) across three analytic approaches. Main (black): full ANCOVA model; Reduced (gray): simplified ANCOVA excluding period and sequence effects; Crude (tan): unadjusted paired comparison. Gray dashed lines show conventional thresholds (0.2, 0.5, 0.8).

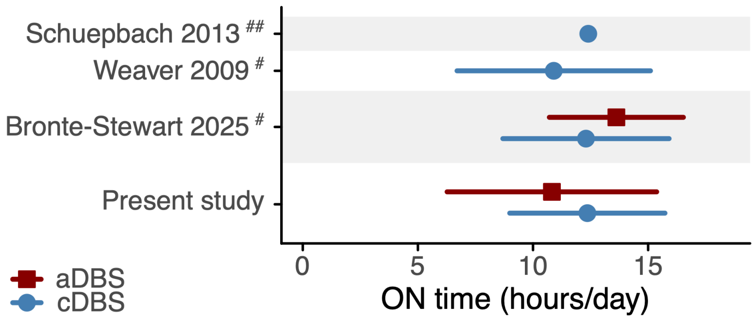


**Supplementary Figure S2. Cross-study comparison of treatment-phase ON time**

Treatment-phase ON time across the present study and selected prior DBS cohorts, all of which were assessed during chronic stimulation under medical therapy. Red squares indicate aDBS and blue circles indicate cDBS. Error bars represent standard deviations. #, ON without troublesome dyskinesia or good ON time[19, 34]; ##, time with good mobility and no dyskinesia[4]. For Bronte-Stewart 2025[19], the aDBS value corresponds to dual-threshold (DT) aDBS. For Schuepbach 2013[4], SD could not be reconstructed from the published report.

**Supplementary References**


S1. Byron Jones MGK. Design and Analysis of Cross-Over Trials. 3rd Edition ed: Routledge; 2014.

S2. Dwan K, Li T, Altman DG, Elbourne D. CONSORT 2010 statement: extension to randomised crossover trials. *BMJ*. 2019;366:l4378.

S3. Kenward MG, Jones B. The analysis of data from 2 x 2 cross-over trials with baseline measurements. *Stat Med*. 1987;6:911-26.

S4. Li B, Zhou Z, Zhang L, Yang Y. Testing treatment-by-period interaction in four-period crossover trials. *Pharm Stat*. 2020;19:145-63.

S5. Willan AR, Pater JL. Using baseline measurements in the two-period crossover clinical trial. *Control Clin Trials*. 1986;7:282-9.

S6. Koo TK, Li MY. A Guideline of Selecting and Reporting Intraclass Correlation Coefficients for Reliability Research. *J Chiropr Med*. 2016;15:155-63.


**Note:** References 4, 19, 20, 25, 26, and 34 are listed in the reference list of the main manuscript.
